# Clinical significance and possible mechanism of different ventricular electrogram morphology in selective left bundle branch pacing

**DOI:** 10.1101/2024.12.02.24318355

**Authors:** Dongjuan Wang, Longfu Jiang, Jiabo Shen, HengDong Li

**Author notes:** **Address for correspondence**: Longfu Jiang, Postal address: 41# northwest street, Ningbo, Zhejiang, China 315010. **Disclosures:** None.

## Abstract

**Background:** Currently, splitting of electrogram (EGM) or electrocardiogram (ECG) under threshold test are used as the gold standard to assess Left bundle branch (LBB) capture in LBB area pacing. However, discrete intracardiac ventricular EGM has not been reported until now. This study aims to explore the clinical significance and possible mechanism of different pacing ventricular EGM morphologies in selective LBB pacing.

**Methods:** Only patients with evidence of selective LBB pacing (splitting of EGM under threshold test) were included. According to the differences between intrinsic and paced ventricular EGM morphologies, the participants were further divided into three groups: concordant EGM (CE) group, similar EGM (SE) group and discordant EGM (DE) group. Baseline characteristics, indications for pacing, pacing parameters, and V6 R-wave peak time were analyzed.

**Results:** 274 patients (85.6%) achieved successful selective LBB pacing. After excluding 34 LBBB patients, LBB potential was recorded in 192 (80%) of 240 patients. In patients with LBB potential, the correlation between V-V6(P) RWPT and V-V6(S) RWPT in CE group (r=0.083, P*<*0.0001) and SE group (r=0.766, P*<*0.0001) were strong. V-V6(S) RWPT was significantly shorter than V-V6(P) RWPT (38.14±9.42 *vs.* 43.68±6.72, *P<0.01*) in DE group. In patients without LBB potential, V-V6(S) RWPT was significantly shorter than V-V6 RWPT (38.14±11.60 vs. 46.15±11.81, P<0.05) in DE group. There was a strong correlation (r=0.943, P<0.0001) between V-V6 RWPT and V-V6(S) RWPT in CE group, a possible correlation (r=0.564, P=0.07) in SE group, while poor correlation (r=0.259, P=0.27) in DE group.

**Conclusion:** The continuous recording technique combined with High Pass-200 Hz filter setting was feasible and effective for confirming selective LBB pacing by discrete EGM. Concordant or similar intrinsic and pacing ventricular EGM indicated that the electric conduction shared the same pathway, while discordant intrinsic and pacing ventricular EGM indicated that the electrical stimulation is conducted through different pathway.

**WHAT IS KNOWN?:** 1. Left bundle branch (LBB) pacing is a novel physiological pacing strategy.

2. Double transition in QRS morphology during threshold testing was considered as the criteria for LBB capture, and splitting of EGM under threshold test was used as the gold standard to assess selective LBB pacing.

3. Identifying discrete local ventricular EGM is still a challenging task.

**WHAT THE STUDY ADDS:** 1. The continuous recording technique combined with High Pass-200 Hz filter setting was feasible and effective for confirming selective LBB pacing by discrete EGM.

2. Different pacing ventricular EGM morphologies compared with intrinsic EGM accounted for clinical significance and possible mechanism: concordant or similar intrinsic and pacing ventricular EGM indicated that the electric conduction shared the same pathway, while discordant intrinsic and pacing ventricular EGM indicated that the electrical stimulation is conducted through different pathway.

3. The anatomical structure of LBB and its fascicular branch was complex, which could not be adequately recorded by 12-lead ECG and EGM.

## Introduction

Left bundle branch (LBB) pacing is a novel physiological pacing strategy, which captures the left conduction system with a low pacing threshold and high R-wave amplitudes ^1^. LBB pacing use is growing fast recently with the advent of tools which have facilitated implantation. Recently, a number of clinical studies have suggested that LBB pacing yielded long-term clinical outcomes superior to those of left ventricular septal (LVS) pacing and biventricular pacing ^2^. LBB pacing activates the normal cardiac conduction and provides ventricular electrical and mechanical synchrony ^3–5^. It’s important to confirm selective LBB capture achieved during LBB pacing, not just the capture of only the adjacent left ventricular myocardium. According to the recent consensus statement on conduction system pacing, double transition in QRS morphology during threshold testing was considered as the criteria for LBB capture, and splitting of EGM under threshold test was used as the gold standard to assess selective LBB pacing ^6^. However, discrete intracardiac ventricular EGM has not been reported until now.

The distribution of left bundle branch is still not fully elucidated, as well as their relationship with V6 R-wave peak time (RWPT). During LBB pacing, the criteria for identifying the lead position need reliable indicators, which may also provide information for whether electrical stimulation conducted through the same or different pathway compared with intrinsic electric conduction pathway. With our continuous technique of LBB pacing, discrete intracardiac ventricular EGM with different morphologies were observed when selective LBB pacing was achieved. This study aims to explore the clinical significance and possible mechanism of different ventricular EGM morphology in selective LBB pacing, which may provide evidence for lead position.

## Methods

### Patient population

This study enrolled consecutive patients underwent successful permanent pacemaker implantation at Ningbo No 2. Hospital from May 2021 to July 2024. All enrolled patients underwent a novel LBB pacing procedure, which used John Jiang’s connecting cable (Xinwell Medical Technology Co., Ltd., Ningbo, Zhejiang, China) for the continuous pacing and recording technique. The study protocol was approved by the Ethics Committee of the Ningbo No 2. Hospital. Written informed consents were obtained from all patients prior to device implantation.

### Implantation procedure and criteria for selective LBB pacing

The detailed implantation procedure has been described previously ^7, 8^. Briefly, the modified connecting cable had a special rotating device with an IS-1 connector port, by which twelve-lead ECG along with EGM from the pacing lead were continuously recorded with an EPS (EP-Workmate, Abbott Laboratories, Chicago, IL, USA). In contrast to the interrupted pacing method, in which the alligator cable is momentarily disconnected during lead rotation, this strategy enables the real-time monitoring of progressive alterations in the paced QRS complex and EGM as the lead penetrates deeper into the interventricular septum. The band-pass filter for the pacing lead was set to “High Pass-200 Hz\Low Pass-500 Hz”. Continuous unipolar pacing at 2V/0.5ms was performed during the whole period of lead implantation and the amplitude was set as 0.5mV/cm. The lead screwing was stopped until the stimulus-V6RWPT for two adjacent paced QRS complexes was abruptly decreased more than 5ms, along with QRS changes such as the new-onset or deeper s-wave in lead V6, or initial r-wave in lead V1 ^9^, then the output was gradually decremented until an smooth interval was observed in intracardiac EGM. If no smooth interval was observed during decremented output, the lead will be screwed with extreme caution under threshold output until the discrete ventricular EGM was found. The discrete local ventricular component and smooth interval with decrementing output during the EGM recording were used as the golden standard of the selective LBB capture.

### Data collection

Baseline characteristics of the patients, indications for pacing, and pacing parameters were recorded at implants. When patients had more than one indication, the dominant indication was used. Unipolar pacing thresholds, R-wave amplitudes, and impedances were recorded. Intracardiac EGM and twelve-lead ECG data were continuously recorded during the whole process form the beginning to the end of lead screwing. The presence of LBB potential was noted.

For patients with LBB potential, the following parameters were recorded: (1) S-V interval, pacing stimulus-ventricular potential interval; (2) P-V interval, LBB potential-ventricular potential interval; (3) S-V6 RWPT, pacing stimulus-V6 R wave peak time; (4) P-V6 RWPT, LBB potential-V6 R wave peak time; (5) V6(S) RWPT, pacing V6 R-wave peak time, which calculated by S-V6 RWPT minus S-V interval; (6) V6(P) RWPT, intrinsic V6 R-wave peak time, which calculated by P-V6 RWPT minus P-V interval.

For patients without LBB potential, the following parameters were recorded: (1) S-V interval, pacing stimulus-ventricular potential interval; (2) S-V6 RWPT, pacing stimulus-V6 R wave peak time; (3) V6(S) RWPT, pacing V6 R-wave peak time, which calculated by S-V6 RWPT minus S-V interval; (4) V6 RWPT, intrinsic V6 R-wave peak time, was measured from the onset of the ventricular EGM to R-wave peak in lead V6.

The analysis of discrete local ventricular EGM morphology was performed using endocardial channel recording with on-screen electronic calipers, at 200 mm/s paper speed. All ventricular EGM morphologies were independently analyzed by two medical practitioners who were highly experienced in EGM interpretation. According to the differences between intrinsic and paced ventricular EGM morphologies, the participants with or without LBB potential were further divided into three groups: (1) Concordant EGM (CE), the paced ventricular EGM morphology was matched to the intrinsic ventricular EGM morphology, and the difference of the main wave amplitude less than or equal to 1/3 of the intrinsic ventricular EGM main wave amplitude; (2) Similar EGM (SE), the paced ventricular EGM morphology was matched to the intrinsic ventricular EGM morphology, and the difference of the main wave amplitude exceeded 1/3 of the intrinsic ventricular EGM main wave amplitude; (3) Discordant EGM (DE), the paced ventricular EGM morphology components were totally different from the intrinsic ventricular EGM. Figure 1 and 2 showed typical cases accepted selective LBB pacing with or without LBB potential respectively. All participants’ EGM ang 12-lead ECG could be found in attachment files.

**Figure 1.**
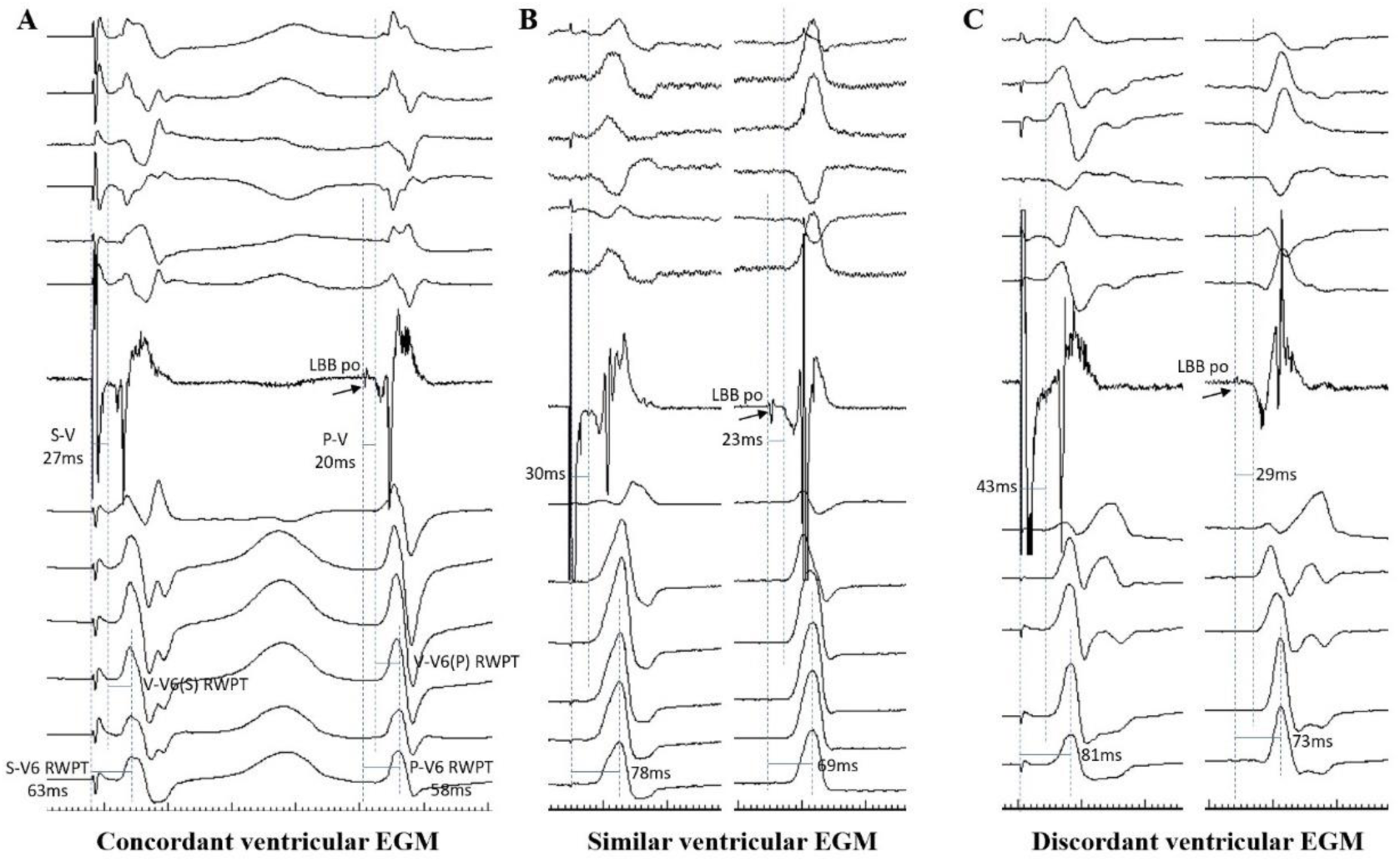
Typical cases accepted LBB pacing with LBB potential. A, Concordant ventricular EGM; B, Similar ventricular EGM; C, Discordant ventricular EGM. LBB po, left bundle branch potential; P-V, LBB potential-ventricular potential interval; S-V, pacing stimulus-ventricular potential interval; P-V6 RWPT, LBB potential-ventricular potential interval; S-V6 RWPT, pacing stimulus-V6 R wave peak time; V-V6(P) RWPT, pacing V6 R-wave peak time; V-V6(S) RWPT, intrinsic V6 R-wave peak time. #P<0.005 compared with CE group.

**Figure 2.**
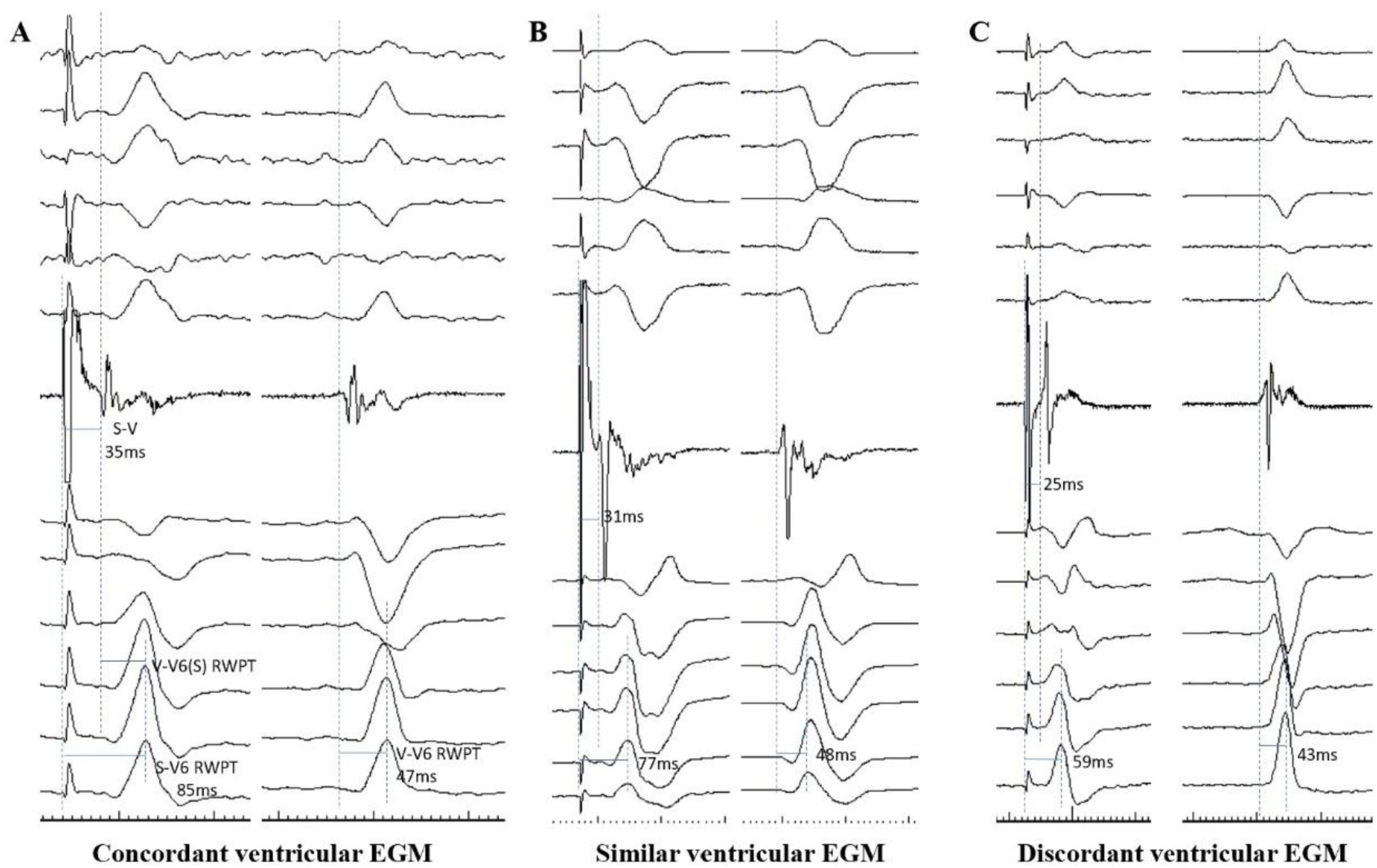
Typical cases accepted LBB pacing without LBB potential. A, Concordant ventricular EGM; B, Similar ventricular EGM; C, Discordant ventricular EGM. S-V, pacing stimulus-ventricular potential interval; S-V6 RWPT, pacing stimulus-V6 R wave peak time; V-V6(S) RWPT, pacing V6 R-wave peak time; V-V6 RWPT, intrinsic V6 R-wave peak time.

### Statistical analysis

Continuous data were expressed as mean values ± standard deviation (SD) and were analyzed using ANOVA followed by a Bonferoni correction for post hoc *t* test. Categorical data were expressed as percentages and were assessed with Chi square and Fisher exact tests. Unequal variances were assessed with the Mann–Whitney U test. Correlation analysis between changes of V-V6(P) RWPT and V-V6(S) RWPT was performed using the Pearson’s method as appropriate for data distribution. Values of *P<0.05* were considered statistically significant. Statistical analysis was performed using GraphPad Prism software version 6.0 (GraphPad Software, USA).

## Results

### Baseline clinical characteristics and pacing characteristics

This was a retrospective, single-center, observational study which included all patients who underwent attempted LBB pacing implant from May 2021 to July 2024. There were 320 patients underwent attempted LBB pacing, 21 patients received left ventricular septal (LVS) pacing because of failing to capture LBB system, and successful LBB pacing was achieved in 299 patients (93.4%) evidenced by splitting of EGM or ECG under threshold test. QRS morphology transition under threshold test. 25 patients were confirmed as having non-selective LBB (NS-LBB) pacing (NS-LBB transform to LVSP), and 274 patients (85.6%) accepted successful selective LBB (S-LBB) pacing (NS-LBB pacing transform to S-LBB pacing) with discrete EGM as endpoint. 34 complete LBB block (LBBB) cases were excluded, and LBB potential was recorded in 192 (80%) of 240 patients (Figure 3).

**Figure 3.**
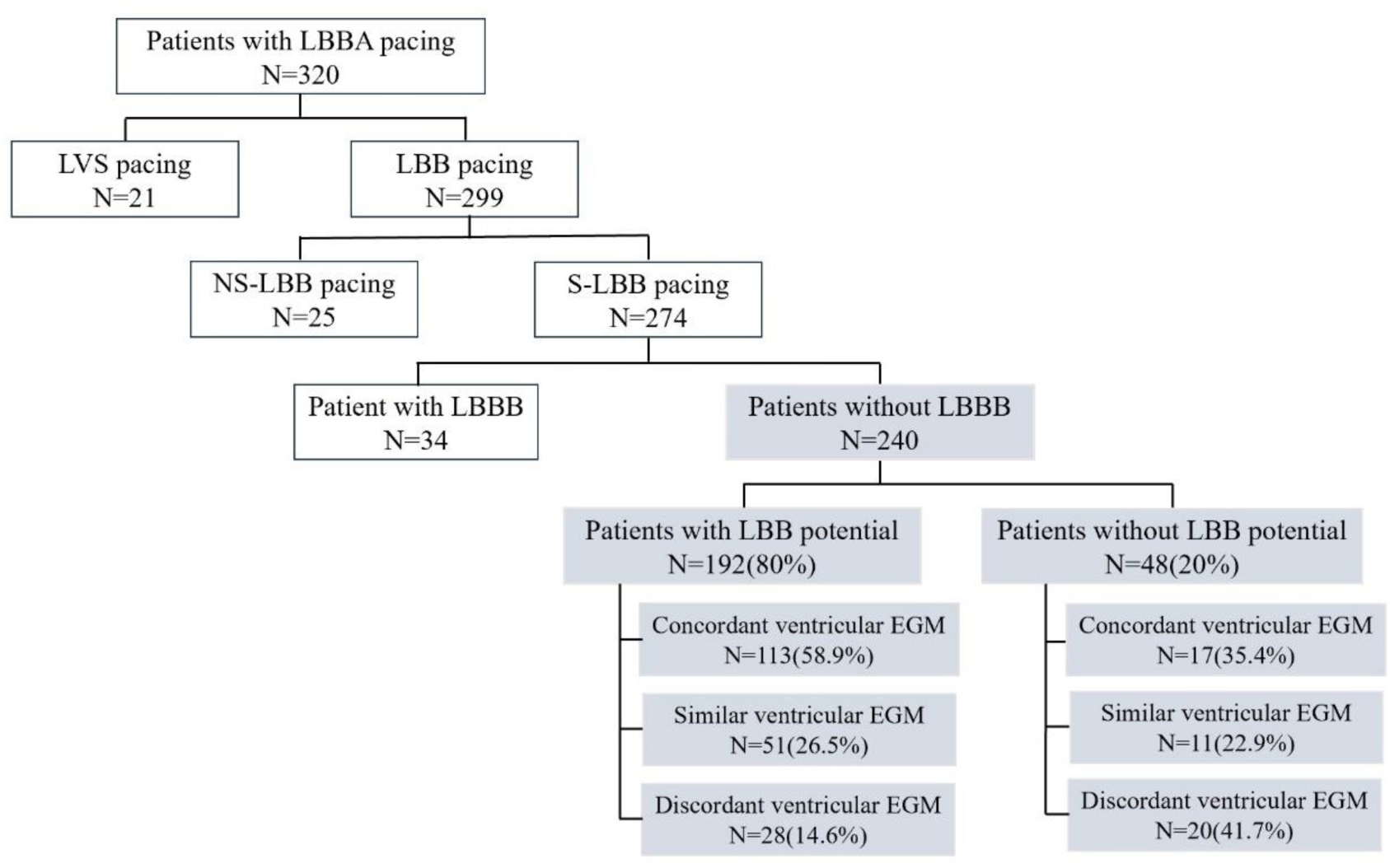
Flowchart for different changes in ventricular EGM of patients receiving LBBAP. LBBA pacing, left bundle branch area pacing; LVS pacing, left ventricular septal pacing; NS-LBB pacing, non-selective left bundle branch pacing; S-LBB pacing, selective left bundle branch pacing; LBBB, left bundle branch block.

Baseline clinical characteristics and pacing characteristics were summarized in Table 1. The mean age was 77.44±9.99 in patients without LBB potential (N-Po), which was significantly older than patients presented with LBB potential (Po, 73.91±8.44, *P<0.05*). 98/192 (51.0%) were male in Po group, and 30/48 were (62.5%) were male in N-Po group. The pacing indications were atrioventricular block (AVB) in 129 (67.2%) patients, and sick sinus syndrome (SSS) in 63 (32.8%) in Po group, while were AVB in 42 (87.5%) patients, and SSS in 6 (12.5%) in N-Po group, which exhibited statistical difference between the two groups (*P<0.01*). We further analyzed the combined arrhythmia, and found that left anterior fascicular block (LAFB) and left posterior fascicular block (LPFB) prevalence was higher in N-Po (35.4% *vs.* 14.6%, *P<0.01*), as well as RBBB (64.6% *vs.* 41.7%, *P<0.01*) compared with Po group. However, there was no significant difference in atrial fibrillation (AF) prevalence (14.6% *vs.* 21.9%, *P>0.05*). The final pacing threshold (unipolar pacing), R-wave amplitude, and impedance were recorded before the pacemaker implantation. The impedance and myocardial threshold were similar between Po and N-Po groups. The LBB threshold was lower in Po group (0.58±0.38 *vs.* 0.80±0.49, *P<0.001*), and R-wave amplitude was lower in N-Po patients (8.88±4.60 *vs.* 10.59±5.25, *P<0.05*). It showed that there were 58.9% (113/192) patients in Po group exhibited concordant ventricular EGM, which was significant higher compared with patients (17/48, 35.4%) in N-Po group (*P<0.01*), while the prevalence of discordant ventricular EGM was statistical lower in patients with LBB potential compared with patients without LBB potential (14.6% *vs.* 41.7%, *P<0.0001*).

**Table 1.**
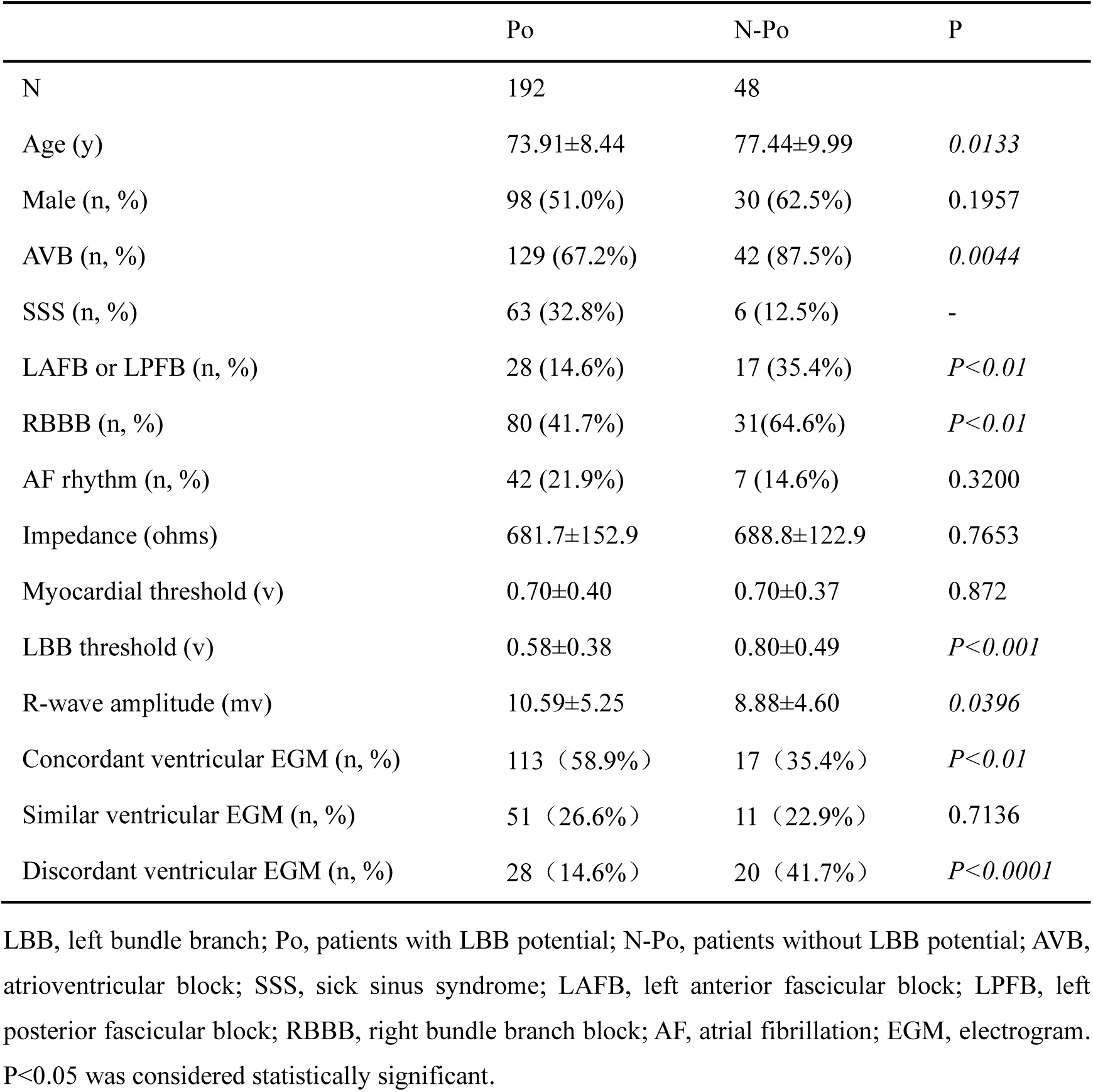
Clinical characteristics of the patients accepted S-LBB pacing.

### The ECG performance of patients with LBB potential

Subgroup analysis of patients with LBB potential demonstrated no significance in P-V interval among the CE, SE and DE groups, as well as S-V interval. We further investigated the difference between the P-V interval and S-V interval within groups. It showed that S-V interval demonstrated a significant increase compared with P-V interval in all three groups, which may be associated with LBB Wenckebach conduction of faster pacing rate than spontaneous rate. P-V6 RWPT and S-V6 RWPT, which indicated LBB potential or pacing stimulus to V6 R wave peak time independently, demonstrated no statistical difference among the three groups. However, S-V6 RWPT were longer compared with P-V6 RWPT in CE and SE group, while there was no difference in DE group (Table 2, Figure 4). The above results suggested that difference liked to left ventricular activation time may exist among the three groups.

**Figure 4.**
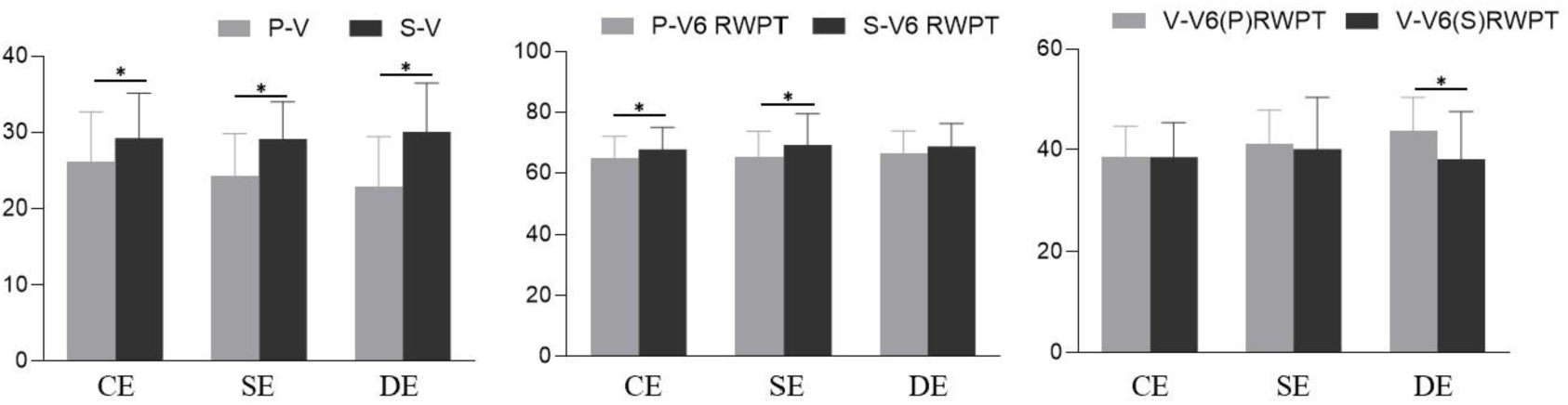
ECG parameters of the patients with LBB potential. CE, concordant EGM; SE, similar EGM; DE, discordant EGM; P-V, LBB potential-ventricular potential interval; S-V, pacing stimulus-ventricular potential interval; P-V6 RWPT, LBB potential-ventricular potential interval; S-V6 RWPT, pacing stimulus-V6 R wave peak time; V-V6(P) RWPT, pacing V6 R-wave peak time; V-V6(S) RWPT, intrinsic V6 R-wave peak time. ***\*****P<0.05*.

**Table 2.** ECG parameters of the patients with LBB potential.

|  | CE | SE | DE |
| --- | --- | --- | --- |
| P-V | 26.06±6.63 | 24.16±5.64 | 22.86±6.54 |
| S-V | 29.22±5.94 | 29.08±4.91 | 30.04±6.40 |
| P-V6 RWPT | 64.73±7.36 | 65.33±8.37 | 66.54±7.33 |
| S-V6 RWPT | 67.77±7.23 | 69.20±10.35 | 68.82±7.45 |
| V-V6(P) RWPT | 38.67±6.02 | 41.16±6.73 | 43.68±6.72<sup>#</sup> |
| V-V6(S) RWPT | 38.55±6.82 | 40.12±10.26 | 38.14±9.42 |

LBB, left bundle branch; CE, concordant EGM; SE, similar EGM; DE, discordant EGM; P-V, LBB potential-ventricular potential interval; S-V, pacing stimulus-ventricular potential interval; P-V6 RWPT, LBB potential-ventricular potential interval; S-V6 RWPT, pacing stimulus-V6 R wave peak time; V- V6(P) RWPT, pacing V6 R-wave peak time; V-V6(S) RWPT, intrinsic V6 R-wave peak time. <sup>#</sup>*P*<0.005 compared with CE group.

To make V-V6 RWPT measurement as accurate as possible, the data was calculated by P-V6 RWPT minus P-V interval or S-V6 RWPT minus S-V interval. V-V6(P) RWPT was longer in DE group compared with CE group (43.68±6.72 *vs.* 38.67±6.02, *P<0.005*), however V-V6(S) RWPT did not exhibited significant difference among CE, SE and DE groups. It showed that V-V6(S) RWPT was significantly shorter than V-V6(P) RWPT (38.14±9.42 *vs.* 43.68±6.72, *P<0.01*) in DE group, but there was no significance between V-V6(P) RWPT and V-V6(S) RWPT in the other two groups. The correlation between V-V6(P) RWPT and V-V6(S) RWPT in CE group (r=0.083, P*<*0.0001) and SE group (r=0.766, P*<*0.0001) were strong, while in DE group was poor (r=0.325, P=0.09) (Figure 5). It is indicated that CE and SE group may share different mechanism from DE group for ventricular EGM morphology.

**Figure 5.**
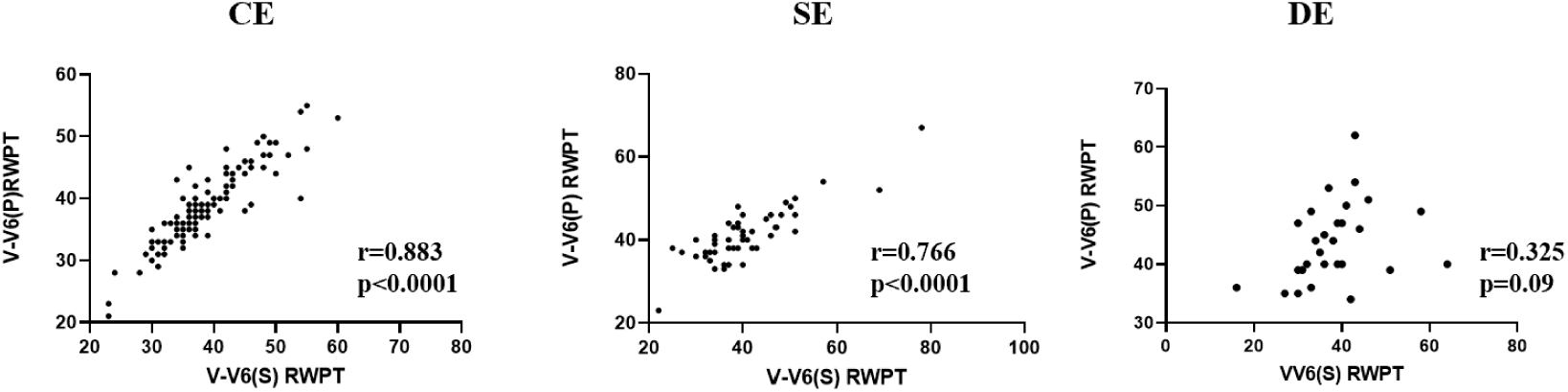
Correlations of V-V6(P) RWPT and V-V6(S) RWPT in patients with LBB potential. CE, concordant EGM; SE, similar EGM; DE, discordant EGM; V-V6(P) RWPT, pacing V6 R-wave peak time; V-V6(S) RWPT, intrinsic V6 R-wave peak time.

### The ECG performance of patients without LBB potential

As listed in Table 3, 48 patients were confirmed as selective LBB pacing during the threshold testing without LBB potential recorded. S-V interval did not differ significantly among the CE, SE and DE groups. These were also no statistical differences at V-V6(S) RWPT and V-V6 RWPT among the three groups. However, it showed that V-V6(S) RWPT was significantly shorter than V-V6 RWPT (38.14±11.60 vs. 46.15±11.81, P<0.05) in DE group, while there was no significance between V-V6 RWPT and V-V6(S) RWPT in the other two groups (Figure 6). The above results suggested that in patient without LBBB, the presence of LBB potential may be not mandatory for successful selective LBB pacing. The correlation between V-V6 RWPT and V-V6(S) RWPT in CE group was strong (r=0.943, P<0.0001), while in DE group (r=0.259, P=0.27) were poor (Figure 7). There was possible correlation between V-V6 RWPT and V-V6(S) RWPT in SE group (r=0.564, P=0.07), and the lack of significant correlation may be due to small sample size (n=11).

**Figure 6.**
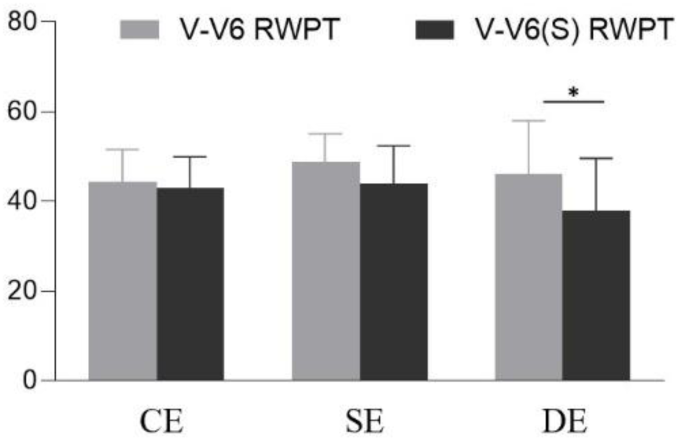
ECG parameters of the patients without LBB potential. CE, concordant EGM; SE, similar EGM; DE, discordant EGM; V-V6(S) RWPT, pacing V6 R-wave peak time; V-V6 RWPT, intrinsic V6 R-wave peak time; ***\*****P<0.05*.

**Figure 7.**
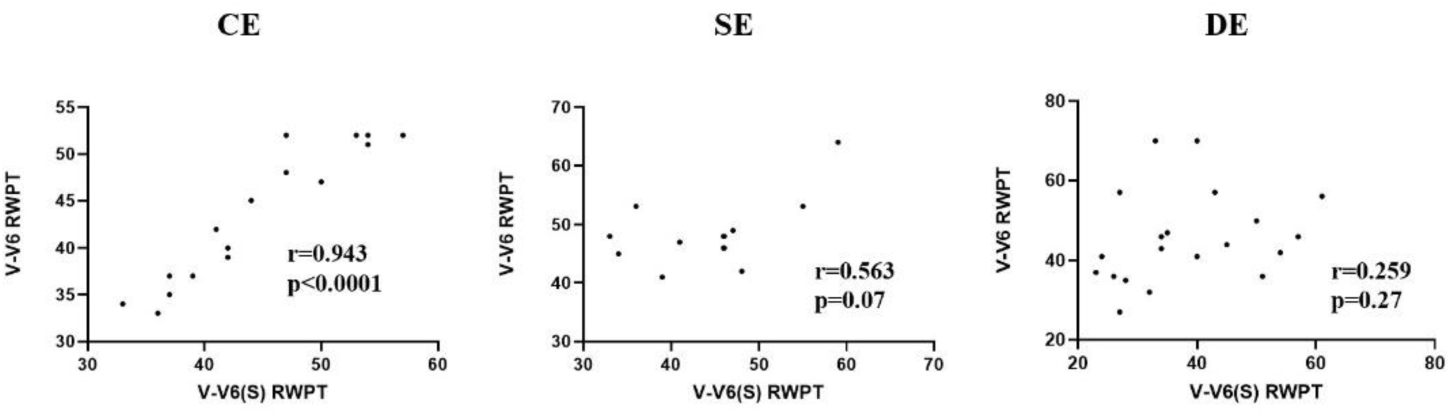
Correlations of V-V6(P) RWPT and V-V6(S) RWPT in patients without LBB potential. CE, concordant EGM; SE, similar EGM; DE, discordant EGM; V-V6(S) RWPT, pacing V6 R-wave peak time; V-V6 RWPT, intrinsic V6 R-wave peak time; ***\*****P<0.05*.

**Table 3.**
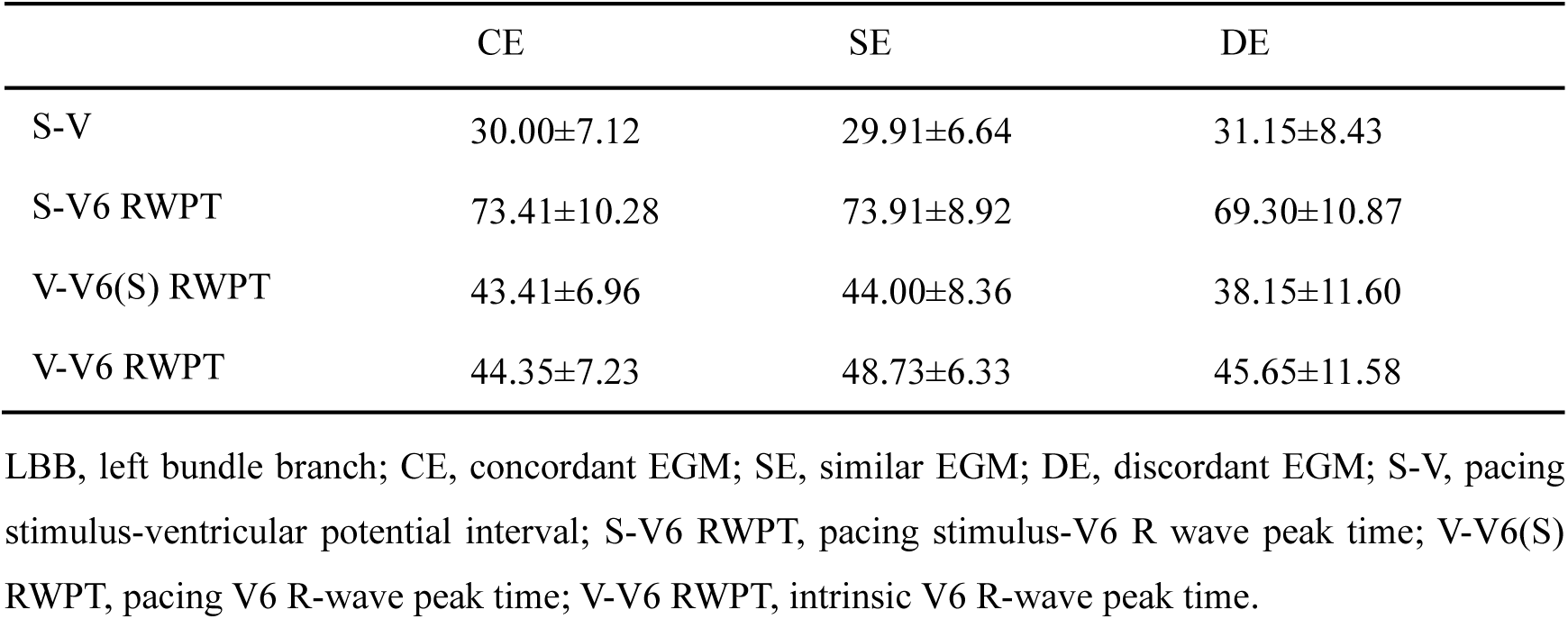
ECG parameters of the patients without LBB potential.

## Discussion

2023 HRS/APHRS/LAHRS guideline on cardiac physiologic pacing suggested that during implantation of conduction system pacing (CSP) leads, it is essential to confirm conduction system capture, which can be challenging ^10^. At present, sensitive and specific criteria for LBB pacing were developed and validated constantly ^11, 12^. European Heart Rhythm Association (EHRA) clinical consensus statement on conduction system pacing implantation have recommended some criteria for confirmation of LBB capture, of which identifying discrete local EGM is crucial for accurately diagnosing selective LBB capture.

However, previous studies have not consistent recommendations on band-pass filter setting for the pacing lead. It suggested that ventricular current of injury (COI) was monitored with minimally filtered signals (High Pass-0.05-0.5 Hz). In clinical practice the high pass filter setting mainly based on the device default settings with 30-40Hz. With this high pass filter, discrete local ventricular EGM may be missed easily because the clipping level limits the display range of large-amplitude endocardial signals. There were also no recommendations on ventricular EGM morphologies during LBB pacing implantation procedures until now. In this study, we used a real-time recording technique with a modified connecting cable, and the band-pass filter for the pacing lead was set to “High Pass-200 Hz\Low Pass-500 Hz”. In general, lower frequencies have a longer wavelength with long-distance transmission. The high pass filter is designed to eliminate unwanted lower frequencies from far filed of ventricle by allowing frequencies higher than the filter settings to pass, which makes the near field higher frequency signals more prominent. During selective LBB pacing, the smooth interval is observed between the pacing artifact and the paced discrete ventricular component, which attributed to only higher frequencies of local ventricle EGM sensed on the tip lead. Demonstration of discrete local EGM may be challenging due to short stimulus to ventricular intervals, effects of stimulus artifact, and far-field recording by the pacing lead. The above band-pass filter may avoid ignoration or misinterpretation subtle EGM changes, and laid the foundation for investigating clinical significance and possible mechanism of different intrinsic and pacing ventricular EGM. Our previous study has reported that discrete local EGM under high Pass-200 Hz had a sensitivity of 100% and specificity of 100% for selective LBB capture ^13^. With our modified connecting cable and band-pass filter setting, 93.4% patients achieved LBB capture, and 85.6% patients achieved selective LBB capture. Discrete intracardiac EGM with different morphologies were observed in selective LBB pacing, with or without the presence of LBB potential.

Su et al. included 117 consecutive patients with successful LBB pacing, and reported that 115 patients (98.3%) had LBB potential recorded ^14^. William Marion et al. found that for successful LBB capture, the LBB potential may precede, coincide or inside the QRS complex ^15^. Shunmuga et al. reported that LBB potential could be recorded in 81.5% patients (203/249) received successful LBB pacing, which exhibited 2 types such as immediately noted high-frequency potential and resurged sharp biphasic potential over few minutes ^16^. Most of the research focused LBB potential presence in left bundle branch area pacing, however there have been no reports of LBB potential in selective LBB pacing so far. After excluding patients with baseline LBBB, our study included 240 patients received successful selective LBB pacing, and there were 192 (80%) patients with clearly noted LBB potential. It showed that in patients with older age or conduction disorders (AVB, LAFB, LPFB, RBBB), LBB potential presence appeared to be low. The mechanism for LBB potential absence partly because the tip lead may be vertical to the LBB potential vector, and partly because the lead was deployed in the distal aspect of the bloke site, which may be not recorded by 12-lead ECG and EGM.

V-V6 RWPT is often used as a surrogate of left ventricular activation time. Discrete local ventricular EGM showed a smooth interval and a steep deflection when selective LBB pacing was achieved. EGM measurements of electrical intervals combined with ECG measurements in this study could subdue the error of V-V6 RWPT. V6(S) RWPT was calculated by S-V6 RWPT minus S-V interval, and V6(P) RWPT was calculated by P-V6 RWPT minus P-V interval. With selective LBB pacing, the V-V6(P) RWPT in intrinsic rhythm should identical with that of the V-V6(S) RWPT during pacing theoretically. We carefully reviewed all participants’ ventricular EGM, and found that there were main wave amplitude or direction differences between intrinsic and paced ventricular EGM morphologies in some cases. CE group, SE group and DE group were divided based on ventricular EGM morphologies, as described in methods. We further explored the clinical significance and its possible mechanism by comparing the morphologies of intrinsic ventricular EGM and discrete local ventricular EGM.

For patients with LBB potential, it showed that there was no significance between V-V6(P) RWPT and V-V6(S) RWPT in CE and SE group, while V-V6(S) RWPT was significantly shorter than V-V6(P) RWPT in DE group. The anatomical structure of LBB may be a correlate of this phenomenon. Intrinsic impulse or pacing stimulus reached the distal end of the His-Purkinje system, excites the apical myocardium, propagates into the tip-pole myocardium, and was then sensed by the tip lead. For patients in CE and SE group, we speculated that the tip lead was deployed in the intrinsic dominated LBB, and the stimulus electric conduction shared the same pathway. Therefore, the paced ventricular EGM morphology was concordant or similar with intrinsic ventricular EGM morphology (Figure 8A-B). For patients in DE group, the tip lead may be deployed in the non-dominant LBB and much closer to the myocardium, which could stimulate the myocardium early and then returning to the sensing parts. The pacing electrical stimulation was conducted through the non-dominant LBB to the myocardium and propagates to the tip lead, which was different from intrinsic impulse (Fig. 8C-D). The above electric mechanism may result in different ventricular EGM and shorter V-V6(S) RWPT in DE group.

**Figure 8.**
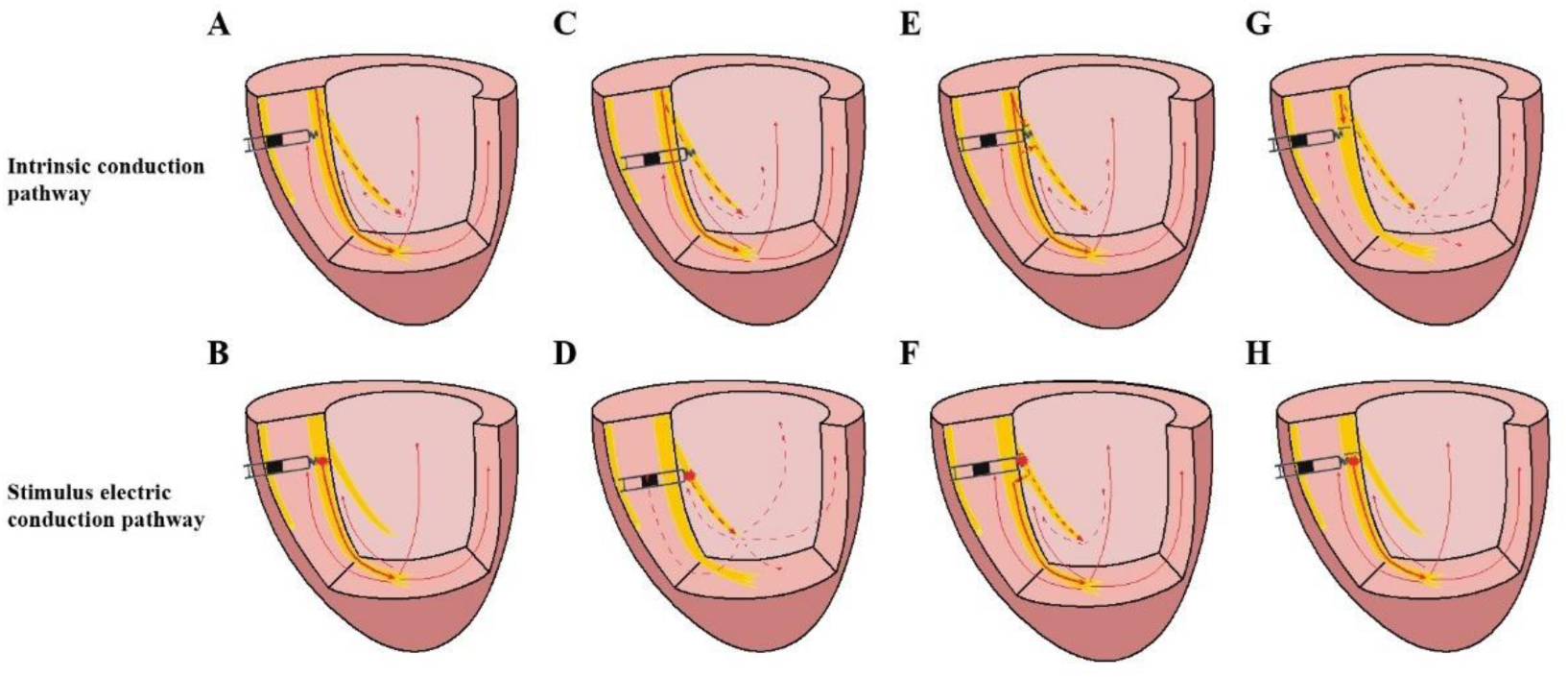
Schematic diagram of different conduction patterns in patient accepted selective LBB pacing. A-B, for patients with LBB potential in CE and SE group, the tip lead was deployed in the intrinsic dominated LBB, and the stimulus electric conduction shared the same pathway; C-D, for patients with LBB potential in DE group, the tip lead may be deployed in the non-dominant LBB, and the electrical stimulation was conducted through the non-dominant LBB and propagates to the tip lead, which was different from intrinsic impulse; E-F, for patients without LBB potential in CE and SE group, the lead tip was located on the distal aspect of the bloke site, but there were functional transverse interconnections within the bundle branches, through which the pacing electrical stimulation conducted to the dominant conduction system; for patients without LBB potential in DE group, there was no functional transverse interconnection, so the pacing electrical stimulation was conducted through the non-dominant LBB to the myocardium.

After excluding patients with baseline LBBB, 48 patents were confirmed as selective LBB pacing with discrete EGM. Among these patients, more than 40% were found with discordant ventricular EGM morphologies, which was significantly higher compared with patients with LBB potential (41.7% vs. 14.6%, P<0.0001). There was no significance between V-V6 RWPT and V-V6(S) RWPT in CE and SE group, while V-V6(S) RWPT was significantly shorter than V-V6 RWPT in DE group. The explanations for the above phenomenon may be considered as follows: (1) The tip lead may be vertical to the LBB potential vector, which was supported by that there was no statistical difference in V-V6 RWPT among the CE, SE and DE groups. But this may be an accidental phenomenon resulted in LBB potential absence. (2) The more common reason was that the deployed site of lead tip was located on the distal aspect of the bloke site, which may be not recorded by 12-lead ECG and EGM. For patients in CE and SE group, the tip lead may be deployed in the non-dominant LBB, but there were functional transverse interconnections within the bundle branches, through which the pacing electrical stimulation conducted to the dominant conduction system (Figure 8E-F). Therefore, identical ventricular EGM morphologies were observed. For patients in DE group, there was no functional transverse interconnection, so the pacing electrical stimulation was conducted through the non-dominant LBB to the myocardium (Figure 8G-H).

We further analysis the difference of V-V6 RWPT in DE group with or without LBB potential. It showed that there was no statistical difference in intrinsic V-V6 RWPT between patients with and without LBB potential (43.68±6.72 *vs.* 45.65±11.58, *P>0.05*), as well as V-V6(S) RWPT (38.14±9.42 *vs* 38.15±11.60, *P>0.05*). In this study, we also found that the intrinsic V-V6 RWPT was significantly higher in patients without LBB potential in CE and SE group (46.07±7.11 *vs.* 39.45±6.34, *P<0.0001*), as well as V-V6(S) RWPT (43.64±7.39 *vs.* 39.04±8.05, *P<0.01*). The above data further confirmed that for patients with LBB potential in CE and SE group, the tip lead was deployed in the intrinsic dominated LBB, and the stimulus electric conduction shared the same pathway, while in other situations the tip lead may be deployed in non-dominant LBB. For patients without LBB potential, longer native V-V6 RWPT indicated slower conduction velocity of Purkinje fibers, which was also supported by higher incidence of fascicular block in patients without LBB potential.

LBB pacing might generate a more physiologic left ventricular activation pattern and better ventricular synchrony characteristics. To the best of our knowledge, no previous study has described the ventricular EGM morphologies with or without the LBB potential in selective LBB pacing. This study demonstrated that although patients in DE group manifested with shorter V-V6(S) RWPT, the electrical synchrony was challenged by different intrinsic and pacing ventricular EGM morphologies. Shorter V-V6(S) RWPT may not predict the physiological synchrony. We speculated that the anatomical structure of LBB was more complex beyond we currently realized. Regarding physiological conduction characteristics and coordination of left ventricular function, concordant or similar intrinsic and pacing ventricular EGM is preferred, but whether it contributed additional clinical benefits in patients still needs further investigation.

### Limitations

It should be admitted that there are still some shortcomings in the research. This study was based on data from one center, and the results might not be universally applicable due to population and implantation technique differences. This was a cross-sectional study without follow up, and additional clinical trials are needed to confirm these findings.

### Conclusions

Understanding the electrophysiological characteristics of the ventricular EGM is important because it may predict a better electrical synchrony. The main findings of our study are as follows: (1) With our modified connecting cable and band-pass filter setting, confirmation of selective LBB pacing by discrete EGM was feasible and effective; (2) The anatomical structure of LBB and its fascicular branch was complex, which could not be adequately recorded by 12-lead ECG and EGM; (3) For patients with or without LBB potential, concordant or similar intrinsic and pacing ventricular EGM indicated that the electric conduction shared the same pathway, while discordant intrinsic and pacing ventricular EGM indicated that the electrical stimulation is conducted through different pathway.

## Data Availability

All data can be viewed in the attachment files.

## Nonstandard Abbreviations and Acronyms

LBBA pacing: left bundle branch area pacing
LBB pacing: left bundle branch pacing
LVS pacing: left ventricular septal pacing
NS-LBB pacing: non-selective left bundle branch pacing
S-LBB pacing: selective left bundle branch pacing
P-V: LBB potential-ventricular potential interval
S-V: pacing stimulus-ventricular potential interval
P-V6 RWPT: LBB potential-ventricular potential interval
S-V6 RWPT: pacing stimulus-V6 R wave peak time
V-V6(P) RWPT: pacing V6 R-wave peak time
V-V6(S) RWPT: intrinsic V6 R-wave peak time
CE: concordant electrogram
SE: similar electrogram
DE: discordant electrogram

## Reference

1. Keene D, Anselme F, Burri H, Perez OC, Curila K, Derndorfer M, Foley P, Geller L, Glikson M, Huybrechts W, Jastrzebski M, Kaczmarek K, Katsouras G, Lyne J, Verdu PP, Restle C, Richter S, Timmer S, Vernooy K and Whinnett Z. Conduction system pacing, a European survey: insights from clinical practice. Europace. 2023;25.

2. Zhu H, Qin C, Du A, Wang Q, He C, Zou F, Li X, Tao J, Wang C, Liu Z, Xue S, Zeng J, Qian Z, Wang Y, Hou X, Ellenbogen KA, Gold MR, Yao Y, Zou J and Fan X. Comparisons of long-term clinical outcomes with left bundle branch pacing, left ventricular septal pacing, and biventricular pacing for cardiac resynchronization therapy. Heart Rhythm. 2024;21:1342–1353.

3. Burri H. Maintaining mechanical synchrony with left bundle branch area pacing. Eur Heart J Cardiovasc Imaging. 2024;25:337–338.

4. Mao Y, Duchenne J, Yang Y, Garweg C, Yang Y, Sheng X, Zhang J, Ye Y, Wang M, Paton MF, Puvrez A, Voros G, Ma M, Fu G and Voigt JU. Left bundle branch pacing better preserves ventricular mechanical synchrony than right ventricular pacing: a two-centre study. Eur Heart J Cardiovasc Imaging. 2024;25:328–336.

5. Vijayaraman P, Subzposh FA, Naperkowski A, Panikkath R, John K, Mascarenhas V, Bauch TD and Huang W. Prospective evaluation of feasibility and electrophysiologic and echocardiographic characteristics of left bundle branch area pacing. Heart Rhythm. 2019;16:1774–1782.

6. Burri H, Jastrzebski M, Cano O, Curila K, de Pooter J, Huang W, Israel C, Joza J, Romero J, Vernooy K, Vijayaraman P, Whinnett Z and Zanon F. EHRA clinical consensus statement on conduction system pacing implantation: endorsed by the Asia Pacific Heart Rhythm Society (APHRS), Canadian Heart Rhythm Society (CHRS), and Latin American Heart Rhythm Society (LAHRS). Europace. 2023;25:1208–1236.

7. Shen J, Jiang L, Cai X, Wu H and Pan L. Left Bundle Branch Pacing Guided by Continuous Pacing Technique That Can Monitor Electrocardiograms and Electrograms in Real Time: A Technical Report. Can J Cardiol. 2022;38:1315–1317.

8. Shen J, Jiang L, Wu H, Cai X, Zhuo S and Pan L. A Continuous Pacing and Recording Technique for Differentiating Left Bundle Branch Pacing From Left Ventricular Septal Pacing: Electrophysiologic Evidence From an Intrapatient-Controlled Study. Can J Cardiol. 2023;39:1–10.

9. Zheng N, Jiang L, Shen J and Zhong J. Guidance on left bundle branch pacing using continuous pacing technique and changes in lead V1 characteristics under real-time monitoring. Front Cardiovasc Med. 2023;10:1195509.

10. Chung MK, Patton KK, Lau CP, Dal Forno ARJ, Al-Khatib SM, Arora V, Birgersdotter-Green UM, Cha YM, Chung EH, Cronin EM, Curtis AB, Cygankiewicz I, Dandamudi G, Dubin AM, Ensch DP, Glotzer TV, Gold MR, Goldberger ZD, Gopinathannair R, Gorodeski EZ, Gutierrez A, Guzman JC, Huang W, Imrey PB, Indik JH, Karim S, Karpawich PP, Khaykin Y, Kiehl EL, Kron J, Kutyifa V, Link MS, Marine JE, Mullens W, Park SJ, Parkash R, Patete MF, Pathak RK, Perona CA, Rickard J, Schoenfeld MH, Seow SC, Shen WK, Shoda M, Singh JP, Slotwiner DJ, Sridhar ARM, Srivatsa UN, Stecker EC, Tanawuttiwat T, Tang WHW, Tapias CA, Tracy CM, Upadhyay GA, Varma N, Vernooy K, Vijayaraman P, Worsnick SA, Zareba W and Zeitler EP. 2023 HRS/APHRS/LAHRS guideline on cardiac physiologic pacing for the avoidance and mitigation of heart failure. Heart Rhythm. 2023;20:e17–e91.

11. Burri H, Jastrzebski M, Cano O, Curila K, de Pooter J, Huang W, Israel C, Joza J, Romero J, Vernooy K, Vijayaraman P, Whinnett Z and Zanon F. EHRA clinical consensus statement on conduction system pacing implantation: executive summary. Endorsed by the Asia-Pacific Heart Rhythm Society (APHRS), Canadian Heart Rhythm Society (CHRS) and Latin-American Heart Rhythm Society (LAHRS). Europace. 2023;25:1237–1248.

12. Li M, Li C, Li J, Yu H, Xu G, Gao Y, Xu B, Sun M, Wang Z, Han Y and Liang Y. An individualized criterion for left bundle branch capture in patients with a narrow QRS complex. Heart Rhythm. 2024;21:294–300.

13. Shen J, Jiang L, Wu H, Li H, Zhang L, Zhong J, Zhuo S and Pan L. High-pass filter settings and the role and mechanism of discrete ventricular electrograms in left bundle branch pacing. Front Cardiovasc Med. 2022;9:1059172.

14. Su L, Xu T, Cai M, Xu L, Vijayaraman P, Sharma PS, Chen X, Zheng R, Wu S and Huang W. Electrophysiological characteristics and clinical values of left bundle branch current of injury in left bundle branch pacing. J Cardiovasc Electrophysiol. 2020;31:834–842.

15. Marion W, Schanz JD, Jr., Patel S, Co ML and Pavri BB. Single operator experience, learning curve, outcomes, and insights gained with conduction system pacing. Pacing Clin Electrophysiol. 2024;47:211–221.

16. Ponnusamy SS, Ganesan V, Ramalingam V, Nachammai P and Vijayaraman P. Electrophysiological characteristics of left bundle branch potential during implantation. Heart Rhythm. 2023;20:1595–1596.

